# Using a Multilingual AI Care Agent to Reduce Disparities in Colorectal Cancer Screening: Higher FIT Test Adoption Among Spanish-Speaking Patients

**DOI:** 10.1101/2024.12.16.24318586

**Authors:** Meenesh Bhimani, R. Hal Baker, Markel Sanz Ausin, Gerald Meixiong, Rae Lasko, Mariska Raglow-Defranco, Alex Miller, Subhabrata Mukherjee, Saad Godil, Anderson Cook, Jonathan D. Agnew, Ashish Atreja

**Affiliations:** Hippocratic AI, Palo Alto, California, USA; WellSpan Health, York, Pennsylvania, USA; School of Population and Public Health, University of British Columbia, Vancouver, BC, Canada; UC Davis Health, Davis, California, USA

**Keywords:** Colorectal cancer screening, artificial intelligence, healthcare disparities, Hispanic Americans, preventive health services

## Abstract

**Background:** Colorectal cancer (CRC) screening rates remain disproportionately low among Hispanic and Latino populations compared to non-Hispanic whites. While artificial intelligence (AI) shows promise in healthcare delivery, concerns exist that AI-based interventions may disadvantage non-English-speaking populations due to biases in development and deployment.

**Objective:** To evaluate the effectiveness of a bilingual AI care agent in engaging Spanish-speaking patients for CRC screening compared to English-speaking patients.

**Methods:** This retrospective analysis examined an AI-powered outreach initiative at WellSpan Health in Pennsylvania and Maryland during September 2024. The study included 1,878 patients (517 Spanish-speaking, 1,361 English-speaking) eligible for CRC screening who lacked active online health profiles. A bilingual AI conversational agent conducted personalized telephone calls in the patient’s preferred language to provide education about CRC screening and facilitate fecal immunochemical test (FIT) kit requests. Primary outcome was FIT test opt-in rate, with secondary outcomes including connect rates and call duration. Statistical analysis included descriptive statistics, bivariate comparisons, and multivariate logistic regression.

**Results:** Spanish-speaking patients demonstrated significantly higher engagement across all measures compared to English-speaking patients: FIT test opt-in rates (18.2% vs. 7.1%, p<0.001), connect rates (69.6% vs. 53.0%, p<0.001), and call duration (6.05 vs. 4.03 minutes, p<0.001). Demographically, Spanish-speaking patients were younger (mean age 57 vs. 61 years, p<0.001) and more likely to be female (49.1% vs. 38.4%, p<0.001). In multivariate analysis, Spanish language preference remained an independent predictor of FIT test opt-in (adjusted OR 2.012, 95% CI 1.340-3.019, p<0.001) after controlling for demographic factors and call duration.

**Conclusions:** AI-powered outreach achieved significantly higher engagement among Spanish-speaking patients, challenging the assumption that technological interventions inherently disadvantage non-English speaking populations. The 2.6-fold higher FIT test opt-in rate among Spanish-speaking patients represents a notable departure from historical patterns of healthcare disparities. These findings suggest that language-concordant AI interactions may help address longstanding disparities in preventive care access. Study limitations include its single healthcare system setting, short duration, and lack of follow-up data on completed screenings. Future research should assess long-term adherence and whether higher engagement translates to improved clinical outcomes.

## Introduction

Colorectal cancer (CRC) screening is crucial for reducing CRC-related mortality by detecting and removing precancerous adenomas or identifying cancer at earlier stages [1], [2]. Various screening methods are available, including colonoscopy, fecal immunochemical tests (FIT), stool DNA tests and CT colonography, each with different levels of effectiveness and potential harms [3]. Recent advances in understanding CRC pathogenesis and improvements in screening technologies have enhanced early detection capabilities.[4] Although some concerns remain regarding the appropriate balance of benefits, risks, and costs, CRC screening is recommended in major guidelines and viewed as a critical public health intervention that can significantly reduce the burden of this prevalent cancer [2], [5], [6], [7].

Despite these benefits, data consistently show lower CRC screening rates among the Hispanic population in the United States compared to non-Hispanic whites, with studies reporting disparities ranging from 13.5% to 17% [8], [9], [10]. Common barriers to CRC screening among Hispanics include fear, cost, lack of awareness, low literacy/education levels, and limited English proficiency [11]. Effective interventions to increase screening rates include culturally tailored patient education, navigation services, and provider training [12], [13], [14]. Nonetheless, disparities persist, with significant regional variations across the United States [10].

Previous research has demonstrated that technological innovations can improve preventive health screening rates among Hispanic populations, suggesting promising applications for artificial intelligence in this domain. Multiple digital approaches have already shown success: mHealth interventions featuring educational videos and interactive multimedia have increased cancer screening participation, while a combination of bilingual patient navigators, secure SMS messaging, and at-home testing achieved an increase in CRC screening rates [15], [16].

Earlier successes with phone-based interventions in Hispanic and psychiatric populations, along with traditional ‘promotora’ programs that improved preventive exam compliance by 35%, suggest that artificial intelligence (AI) could effectively augment these established approaches [17], [18], [19]. Given that tailored patient-centered technologies have consistently improved health outcomes and reduced disparities, AI represents a potential next step in this technological evolution [20].

Nonetheless, its impact in healthcare remains unclear, especially with respect to AI care agents. Indeed, some have suggested that AI-based interventions in healthcare may disadvantage non-English-speaking and minority populations due to biases in development and deployment [21], [22]. These concerns about AI bias in healthcare are well-documented in the literature. Multiple studies have identified systematic disparities in AI algorithm performance across racial and ethnic groups. For example, machine learning models have demonstrated higher predictive accuracy for White patients compared to minority groups across various applications [23]. In health risk prediction, algorithms have been shown to underestimate the care needs of Black patients [24], while in medical imaging, AI systems consistently underdiagnose conditions in underserved populations [25]. These disparities stem from several mechanisms, including underrepresentation of minorities in training datasets, biased feature selection, and implementation contexts that amplify existing healthcare inequities [26], [27]. Given this evidence, careful evaluation of AI-powered interventions specifically targeting minority populations is essential to determine whether they reduce or potentially exacerbate existing healthcare disparities.

Implementation science is considered a key factor in determining the optimal outcome of AI-based initiatives [28]. Implementation science frameworks addressing health equity can provide important theoretical grounding for AI-powered healthcare interventions. The Health Equity Implementation

Framework, which integrates implementation science with healthcare disparities research, identifies multi-level factors affecting implementation from individual patient characteristics to societal contexts.[29] This framework emphasizes culturally relevant factors, clinical encounters, and societal determinants that can influence health equity outcomes. Similarly, the EquIR (Equity-based Implementation Research) framework links pre- and post-implementation population health status with specific equity considerations.[30] These theoretical frameworks suggest that properly designed AI-powered interventions could potentially help address disparities rather than exacerbate them, particularly when they incorporate elements of cultural specificity, language concordance, and responsiveness to community needs. However, implementation science also highlights the importance of evaluating such interventions for their actual impact on health equity outcomes rather than assuming beneficial effects.[31]

This study aims to evaluate whether a multilingual AI care agent can effectively engage Spanish-speaking patients in colorectal cancer screening compared to English-speaking patients. Specifically, we examine whether AI-powered outreach results in comparable or higher FIT test opt-in rates among Spanish-speaking patients, potentially challenging the assumption that technological interventions inherently disadvantage non-English speaking populations. We hypothesize that a language-concordant AI care agent will achieve engagement levels among Spanish-speaking patients that meet or exceed those observed in English-speaking patients, potentially helping to reduce the persistent screening disparities that conventional approaches have struggled to address.

## Methods

We hypothesized that AI-powered outreach using a bilingual AI care agent would demonstrate comparable or higher engagement and FIT test opt-in rates among Spanish-speaking patients compared to English-speaking patients, potentially helping to reduce existing screening disparities between these populations. Our primary aim was to evaluate the effectiveness of a multilingual AI-powered outreach system in improving CRC screening rates among Spanish-speaking patients compared to English-speaking patients.

### Study Design

This retrospective observational study analyzed data from an AI-powered outreach initiative conducted during the week of September 16-24, 2024, at WellSpan Health, an integrated health system serving central Pennsylvania and northern Maryland. The study focused on comparing the effectiveness of multilingual AI-powered outreach for CRC screening between Spanish- and English-speaking patients. As the study analyzed anonymized administrative data, it was exempt from ethics review, and no individual patient consent was required.

### Patient Population

The study population comprised 1,878 patients eligible for CRC screening who did not have active online health system profiles. Of these, 517 (28%) were Spanish-speaking, and 1,361 (72%) were English-speaking patients. Patients were included if they were due for CRC screening and had a verified date of birth. The study specifically targeted patients who were not actively engaged with the healthcare system and were unlikely to be reached through traditional outreach methods such as email.

### Intervention

The intervention used an AI-powered conversational agent named Ana, designed to conduct personalized outreach calls for CRC screening. The platform was developed to engage patients in natural, empathetic telephone conversations while providing education about CRC screening and facilitating FIT test kit requests. The AI care agent was programmed with full bilingual capabilities in English and Spanish, allowing for natural conversation in the patient’s preferred language. The system was designed to engage in culturally appropriate dialogue, explain the importance of CRC screening, address patient questions and concerns, and facilitate the scheduling of FIT test kit deliveries. Calls were initiated to patients’ registered phone numbers, with language preference determined at the start of each interaction by the AI care agent [32]. We used process models to map out the processes required to integrate AI into clinical workflow; identify potential issues before they become barriers in the patient outreach; and adjust strategies for better outcomes [28].

The AI care agent, Ana, is powered by Polaris 3.0, a constellation system designed for real-time multilingual phone conversations. The system incorporates language-specific preprocessing to ensure correct pronunciation of medical terminology and medication dosages, robust error-handling for speech recognition challenges (particularly when patients code-switch between languages), and culturally appropriate dialogue frameworks. The AI agent was trained to detect when patients needed clarification about medical concepts and could adjust its communication style based on patient responses. When discussing colorectal cancer screening, Ana could explain the FIT test procedure, address common concerns, and capture patient preferences while maintaining natural conversation flow in both English and Spanish.

### Outcome Measures

Data were collected automatically through the AI platform’s integrated tracking system. The primary outcome measure was the FIT test opt-in rate, defined as the percentage of patients who agreed to receive a FIT test kit during the AI-powered conversation. Secondary outcomes included connect rate (percentage of successful connections with patients) and call duration.

### Statistical Analysis

Analyses were performed using SPSS version 28 [33]. Descriptive statistics were calculated for demographic characteristics and outcome measures. Differences between Spanish- and English-speaking groups were assessed using independent samples t-tests for continuous variables and chi-square tests for categorical variables. A logistic regression analysis was conducted to evaluate factors associated with FIT test opt-in rates, controlling for age, gender, geographic location, and call duration. The model’s goodness of fit was assessed using the Hosmer-Lemeshow test, and model performance was evaluated using Nagelkerke R^2^. Statistical significance was set at p < 0.001. No missing data were present in the analyzed dataset.

## Results

### Baseline Characteristics

A total of 1,878 patients were included in the analysis, comprising 517 Spanish-speaking (27.5%) and 1,361 English-speaking (72.5%) individuals (Table 1). Significant demographic differences were observed between the language groups. Spanish-speaking patients were younger (mean age 57 ± 8.5 years vs. 61 ± 8.7 years, p<0.001) and more likely to be female (49.1% vs. 38.4%, p<0.001) compared to English-speaking patients. The majority of patients in both groups resided in Pennsylvania, though this proportion was higher among Spanish-speaking patients (99.3% vs. 95.6%, p<0.001).

**Table 1.** Baseline Characteristics by Language Group.

| Characteristic | Spanish-Speaking<br>(n=517) | English-Speaking<br>(n=1,361) | P-value |
| --- | --- | --- | --- |
| **Age, years (SD)** | 57 (8.5) | 61 (8.7) | <.001* |
| **Gender, % (n)** |  |  |  |
| - Female | 49.1% (n=254) | 38.4% (n=523) | <.001** |
| - Male | 50.9% (n=263) | 61.6% (n=838) |  |
| **Geographic Location, % (n)** |  |  |  |
| - Pennsylvania | 99.3% (n=513) | 95.6% (1301) | <.001** |
| - Maryland | 0.6% (n=3) | 3.5% (n=48) |  |
| - Other | 0.2% (n=1) | 0.9% (n=12) |  |

\*Independent samples t-test. \*\*Chi-square test. SD = standard deviation

### Primary and Secondary Outcomes

Spanish-speaking patients demonstrated significantly higher engagement with the AI-powered outreach compared to English-speaking patients across all measured outcomes (Table 2). The FIT test opt-in rate was more than twice as high among Spanish-speaking patients (18.2% vs. 7.1%, p<0.001). Similarly, connect rates were substantially higher in the Spanish-speaking group (69.6% vs. 53.0%, p<0.001).

**Table 2:**
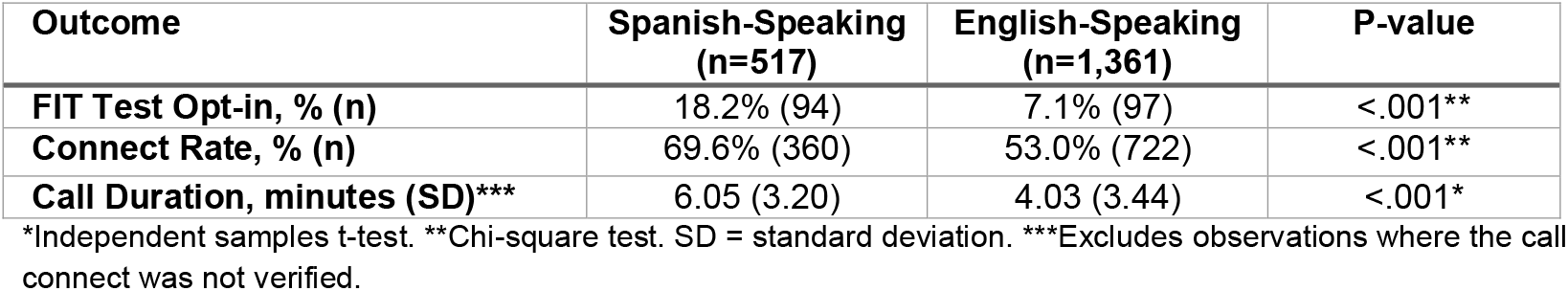
Outcomes by Language Group.

| Outcome | Spanish-Speaking<br>(n=517) | English-Speaking<br>(n=1,361) | P-value |
| --- | --- | --- | --- |
| **FIT Test Opt-in, % (n)** | 18.2% (94) | 7.1% (97) | <.001** |
| **Connect Rate, % (n)** | 69.6% (360) | 53.0% (722) | <.001** |
| **Call Duration, minutes (SD)\*\*\*** | 6.05 (3.20) | 4.03 (3.44) | <.001* |

\*Independent samples t-test. \*\*Chi-square test. SD = standard deviation. \*\*\*Excludes observations where the call connect was not verified.

Additionally, Spanish-speaking patients engaged in longer conversations with the AI care agent (6.05 minutes vs. 4.03 minutes, p<0.001).

### Multivariate Analysis

A logistic regression analysis revealed that Spanish language preference remained an independent predictor of FIT test opt-in after adjusting for age, gender, geographic location, and call duration (Table 3). Spanish-speaking patients were more than twice as likely to opt in for FIT testing compared to English-speaking patients (adjusted OR 2.012, 95% CI 1.340-3.019, p<0.001). Call duration was also significantly associated with opt-in likelihood (adjusted OR 1.682 per minute, 95% CI 1.583-1.787, p<0.001). Age, gender, and geographic location were not significantly associated with FIT test opt-in rates.

**Table 3.** Logistic Regression Analysis of Factors Associated with FIT Test Opt-in.

| Variable | B Coefficient (SE) | Adjusted OR (95% CI) | P-value |
| --- | --- | --- | --- |
| **Age (per year)** | -0.010 (0.11) | 0.990 (0.968 – 1.1012) | N/S |
| **State** |  |  |  |
| - Pennsylvania | Reference | 1.00 | -- |
| - Maryland or Other | -1.527 (0.908) | 0.217 (0.037 – 1.288) | N/S |
| **Gender** |  |  |  |
| - Male | Reference | 1.00 | -- |
| - Female | -0.296 (0.200) | 0.744 (0.503 – 1.101) | N/S |
| **Call Duration (per minute)** | 0.520 (0.031) | 1.682 (1.583 – 1.787) | <0.001 |
| **Language** |  |  |  |
| - English | Reference | 1.00 | -- |
| - Spanish | 0.699 (0.207) | 2.012 (1.340 – 3.019) | <0.001 |
| **Constant** | -3.195 (0.715) | -- | <0.001 |

OR = Odds Ratio; CI = Confidence Interval; SE = Standard Error. N/S = not significant. The dependent variable was FIT test opt-in (yes/no). Model fit statistics: Hosmer-Lemeshow $\chi^2(8) = 30.486$ , $p = 0.001$ ; Nagelkerke $R^2 = 0.445$ ; Model $\chi^2(5) = 452.681$ , $p = <0.001$ .

The regression model demonstrated good explanatory power, explaining nearly 445% of the observed variation in FIT test opt-ins (Nagelkerke R^2^ = 0.445). The model demonstrated good overall significance as well (Model χ^2^(5) = 452.681, p<0.001), although the Hosmer-Lemeshow test suggested potential issues with model fit (χ^2^(8) = 30.486, p = 0.001). These findings indicate that language preference was a robust predictor of engagement with CRC screening outreach, independent of other demographic factors.

## Discussion

Our study suggests that AI-powered outreach can effectively engage Spanish-speaking populations in CRC, with outcomes exceeding those observed in English-speaking patients. The 2.6-fold higher FIT test opt-in rate among Spanish-speaking patients (18.2% vs. 7.1%) represents a departure from historical patterns of healthcare disparities – a noteworthy finding given that previous studies have documented consistently lower CRC screening rates among Hispanic and Latino populations (53.4%) compared to non-Hispanic whites (70.4%).[10] Moreover, higher engagement levels observed among Spanish-speaking patients, evidenced by superior connect rates (69.6% vs. 53.0%) and longer call durations (6.05 vs. 4.03 minutes), suggest that AI outreach may help overcome traditional barriers to healthcare access and engagement.

Our multivariate analysis revealed that Spanish language preference remained a significant and independent predictor of FIT test opt-in even after controlling for demographic factors and call duration, with Spanish-speaking patients twice as likely to opt-in to screening compared to their English-speaking counterparts. Our findings that AI-powered outreach achieved higher engagement among Spanish-speaking patients appear to run counter to the substantial body of evidence documenting AI bias in healthcare. Multiple studies have demonstrated that AI applications can disadvantage minority populations through various mechanisms. Obermeyer et al. found that health risk prediction algorithms significantly underestimated the care needs of Black patients, potentially reducing their access to additional care support [24]. Similarly, Embí reported that AI models predicting postpartum depression assigned White patients twice the likelihood of diagnosis compared to Black patients [34]. In medical imaging, Seyyed-Kalantari et al. documented that chest X-ray classifiers consistently underdiagnosed conditions in underserved patient populations [25].

The contrast between these documented disparities and our results warrants careful consideration. Two factors may explain this divergence. First, the nature of our intervention—direct patient outreach and education—differs fundamentally from clinical decision support or diagnostic applications where algorithmic biases have been most extensively documented. Second, the apparent success in Spanish-speaking engagement may reflect the particular receptiveness of this population to personalized outreach addressing a known screening disparity, rather than inherent advantages in the AI methodology itself.

The study benefits from several strengths, including a large sample size with substantial representation of Spanish-speaking patients and complete data capture through automated systems. The real-world implementation in a diverse healthcare setting, coupled with statistical analysis controlling for potential confounders, provides evidence for the effectiveness of the intervention. The direct comparison of language groups within the same intervention period further strengthens our findings.

However, important limitations must be considered. The study’s confinement to a single healthcare system in central Pennsylvania and northern Maryland represents a significant limitation that may affect generalizability to other geographic regions, patient populations, and healthcare delivery models.

Healthcare systems differ considerably in their organizational structures, patient demographics, and existing approaches to preventive care, which may influence the effectiveness of AI-powered interventions across different settings. Additionally, the short study duration may not capture longer-term engagement patterns. A critical limitation is the absence of follow-up data on FIT test completion rates, which prevents assessment of whether higher engagement and opt-in rates actually translated to completed screening tests. This gap leaves an important open question about the intervention’s true impact on screening compliance and its potential to reduce disparities in completed screenings. Our study specifically targeted patients without active online health system profiles, which intentionally focused on less digitally engaged individuals but consequently may have excluded patients who regularly interact with healthcare services through digital platforms. This methodological choice could introduce selection bias. We were also unable to assess socioeconomic, medical, and other factors that might influence engagement, further limiting our ability to fully contextualize our findings. For example, other unaccounted-for factors, such as cultural or social factors among the Spanish-speaking population, might have contributed to higher response rate among this population relative to the English-speaking population.

Our findings suggest that AI-powered outreach can effectively complement existing care delivery systems, particularly for traditionally underserved populations. The higher engagement rates among Spanish-speaking patients indicate that language-concordant AI interactions may help address longstanding disparities in preventive care access and utilization. Healthcare systems seeking to improve screening rates among diverse populations should consider implementing multilingual AI outreach as part of their comprehensive screening strategy.

The promising results of this intervention suggest that technological solutions may not inherently exacerbate healthcare disparities as some have feared. While our findings show higher engagement and FIT test opt-in rates among Spanish-speaking patients, these are preliminary outcomes that represent only the initial steps in the screening process. Nevertheless, these early results have potentially important implications for healthcare policy and resource allocation for technological innovations in healthcare delivery, warranting further investigation of AI-powered interventions across the complete screening continuum.

The markedly higher engagement among Spanish-speaking patients suggests that AI-powered outreach may be particularly effective in reaching traditionally underserved populations. This finding is especially relevant given the historical challenges in engaging Hispanic and Latino communities in preventive care programs. The success of this intervention demonstrates that AI, when properly implemented, can serve as a tool for promoting health equity rather than perpetuating disparities. This approach could be further adapted for other minority populations by incorporating additional languages (such as Mandarin, Vietnamese, or Arabic) and culturally specific communication patterns. Adaptation would require not only linguistic translation but also cultural tailoring of messaging, addressing population-specific barriers to screening, and incorporating community input into AI design. Similar to how the Spanish-language AI agent effectively engaged Hispanic patients, culturally responsive AI systems could potentially bridge engagement gaps for other underserved groups.

Future research should prioritize the assessment of long-term patient adherence to screening recommendations following AI-powered outreach, tracking not only initial engagement but also subsequent screening behaviors over multiple years. Studies should also evaluate the cost-effectiveness of AI interventions compared to traditional outreach methods, including analyses of implementation costs, healthcare resource utilization, and potential cost savings from earlier cancer detection. Most critically, research must determine whether the higher engagement observed with AI-driven interventions ultimately translates to improved clinical outcomes, including increased rates of early-stage cancer detection, reduced cancer mortality, and narrowed disparities in health outcomes. Additionally, future studies should explore how AI outreach models can be specifically tailored to address the unique cultural, linguistic, and health-belief characteristics of diverse ethnic groups, particularly focusing on populations whose primary language is not English. Such culturally adaptive AI approaches might incorporate cultural nuances, dialect variations, and community-specific health concerns to enhance relevance and effectiveness across different populations. Investigation of AI outreach effectiveness in other languages and cultural contexts would provide valuable insights for expansion, while studies examining patient experience with AI-powered interactions would further inform implementation strategies.

## Conclusion

This study demonstrates that carefully designed AI-powered outreach through multilingual AI care agents can effectively engage diverse populations in preventive care, particularly benefiting traditionally underserved Spanish-speaking communities. The significantly higher engagement and FIT test opt-in rates among Spanish-speaking patients challenge previous assumptions about technological interventions potentially disadvantaging non-English speaking populations. These findings suggest that language-concordant AI interactions may help address longstanding disparities in preventive care access.

Future research should prioritize immediate needs to track FIT test completion rates following AI outreach, expand evaluation to diverse healthcare settings beyond a single system, and conduct qualitative research with Spanish-speaking patients to understand engagement factors. In the mid-term, researchers should evaluate cost-effectiveness of multilingual AI interventions compared to traditional methods, including applications beyond CRC screening; extend linguistic capabilities to additional languages with appropriate cultural adaptations; and develop standardized implementation frameworks for equitable deployment. Long-term research priorities should focus on tracking clinical outcomes including early cancer detection rates and mortality to determine if AI outreach ultimately reduces disparities. Investigating how AI care agents can address intersecting social determinants of health contributing to screening disparities, and developing integrated models that combine AI outreach with human navigation for complex cases, may prove particularly fruitful.

As healthcare systems increasingly adopt technological solutions, this research agenda will ensure that AI applications are developed, deployed, and evaluated with explicit attention to health equity principles, including cultural competency, language access, and community engagement. By following this roadmap, future innovations can build on our findings to promote rather than hinder health equity in preventive care and beyond.

## Data Availability Statement

Data supporting the results can be accessed by contacting the corresponding author.

## Ethics Statement

This study analyzed anonymized administrative data that were exempt from ethics review. No individual patient consent was required as all data were de-identified prior to analysis. The study was conducted in accordance with relevant privacy and data protection regulations.

## Declaration of AI Tool Usage

AI tools were used in preparation of this manuscript for initial draft assistance with manuscript language and structure, grammar and style refinement, and reference formatting. All AI-generated content was thoroughly reviewed, verified, and edited by the authors. All statistical analyses, interpretations, and conclusions were conducted and validated by the human authors. The authors take full responsibility for the final content of this manuscript.

